# Lack of Antiviral Activity of Darunavir against SARS-CoV-2

**DOI:** 10.1101/2020.04.03.20052548

**Authors:** Sandra De Meyer, Denisa Bojkova, Jindrich Cinatl, Ellen Van Damme, Christophe Buyck, Marnix Van Loock, Brian Woodfall, Sandra Ciesek

## Abstract

Given the high need and the absence of specific antivirals for treatment of COVID-19 (the disease caused by severe acute respiratory syndrome-associated coronavirus-2 [SARS-CoV-2]), human immunodeficiency virus (HIV) protease inhibitors are being considered as therapeutic alternatives. Prezcobix/Rezolsta is a fixed-dose combination of 800 mg of the HIV protease inhibitor darunavir (DRV) and 150 mg cobicistat, a CYP3A4 inhibitor, which is indicated in combination with other antiretroviral agents for the treatment of HIV infection. There are currently no definitive data on the safety and efficacy of DRV/cobicistat for treatment of COVID-19. The *in vitro* antiviral activity of darunavir against a clinical isolate from a patient infected with SARS-CoV-2 was assessed. DRV showed no activity against SARS-CoV-2 at clinically relevant concentrations (EC_50_ >100 μM). Remdesivir, used as a positive control, showed potent antiviral activity (EC_50_ = 0.38 μM). Overall, the data do not support the use of DRV for treatment of COVID-19.

## Introduction

In December 2019, the severe acute respiratory syndrome-associated coronavirus disease-2 2019 (SARS-CoV-2; COVID-19) emerged in Wuhan, Hubei Province, China (*1*). The virus was subsequently identified as a coronavirus (CoV), in addition to SARS-CoV-1 and Middle East respiratory syndrome CoV (MERS-CoV) that passed from animals to humans where it can cause severe respiratory illness (*2*). As of March 2020, COVID-19 has spread around the world with the WHO declaring a global pandemic (*3*). Given the extent of the COVID-19 pandemic, there is an urgent need to identify potential treatments for the disease as well as to develop a vaccine.

As no specific antivirals for treatment of COVID-19 are available, one avenue of clinical interest is the use of human immunodeficiency virus (HIV) protease inhibitors (PIs) as a therapeutic intervention. The potential for HIV PIs as a treatment for COVID-19 is mainly based on limited virologic and clinical data on the HIV protease inhibitor lopinavir with low-dose ritonavir (as a pharmacoenhancer; LPV/r) in patients infected with severe acute respiratory syndrome related to a coronavirus (SARS-CoV) (*4*). After demonstrating the *in vitro* antiviral activity of LPV against SARS-CoV-1, the clinical response of patients with SARS to a combination of LPV/r and ribavirin was examined. Patients treated with LPV/r had lower rates of adverse clinical outcomes at day 21 following the onset of symptoms compared with historical controls (*4*). However, recent data in hospitalized adults with severe confirmed COVID-19 treated with LPV/r in addition to a standard care of ventilation, oxygen, vasopressor support, antibiotics and renal-replacement therapy showed that there was no significant improvement in time to clinical improvement or mortality at day 28 compared with the standard care (*5*).

The HIV PI darunavir with cobicistat as a pharmacoenhancer (DRV/c, 800/150 mg given orally once daily with food) in combination with other antiretroviral agents is approved for both treatment-naïve and -experienced patients with HIV-1 infection (*6-7*). The efficacy and safety profile of boosted-DRV combination therapy is well-established in the HIV setting, based on phase III clinical studies as well as real-world evidence (*8–11*).

To date, no clear clinical evidence supports the use of DRV (boosted with either ritonavir or cobicistat) in viral diseases other than HIV.

In this paper, the antiviral activity of DRV against SARS-CoV-2 was investigated in an *in vitro* model at clinically relevant concentrations. When DRV/c is taken at the indicated once-daily dose, the median trough plasma concentration of DRV was 3.4 µM (1875 ng/mL). (*6*). This cell culture assay was shown to be suitable for antiviral assays. Productive viral infection takes place in this model with the number of SARS-CoV RNA molecules increasing continuously after infection, indicating that the virus undergoes full replicatory cycles (*12*). Remdesivir (GS-5734), a nucleotide analog initially developed for Ebola virus disease, has shown to inhibit SARS-CoV-2 replication *in vitro* with an EC_50_ equal to 0,770 µM (*13*) and was therefore used as a positive control.

## Methods

### Cell Culture and Virus Preparation

Human colon carcinoma cell line (Caco-2) cells (obtained from the Deutsche Sammlung von Mikroorganismen und Zellkulturen, Braunschweig, Germany) were cultured in Minimal Essential Medium (MEM) supplemented with 10% fetal bovine serum (FBS) and containing penicillin (100 IU/mL) and streptomycin (100 μg/mL) in a 5% CO_2_ atmosphere at 37°C. All culture reagents were purchased from Sigma (Hamburg, Germany).

SARS-CoV-2 was isolated from human samples and cultured in Caco-2 cells, as previously described (*3*). After one passage in Caco-2 cells, viral stocks were stored at –80°C prior to use.

### Assessment of Antiviral Activity by Inhibition of Virus-Induced Cytopathogenic Effect

Confluent layers of Caco-2 cells were cultured at 37°C in a 5% CO_2_ atmosphere for 72 hours on 96 multi-well plates (50,000 cells/well). Cells were challenged with SARS-CoV-2 at a multiplicity of infection (MOI) of 0.01. Virus was added together with the compounds under investigation and incubated in MEM supplemented with 1% FBS.

DRV and remdesivir were synthesized at Johnson & Johnson. To assess *in vitro* antiviral activity, DRV and remdesivir, diluted in MEM without FBS, were added in 4-fold dilutions to a concentration range of 0.02 μM to 100 μM. Cells were then incubated for 48 hours before the cytopathogenic effect (CPE) was visually scored by two independent laboratory technicians. Evaluation of CPE was also done using an 3-(4,5-dimetiltiazol-2-il)-2,5-difeniltetrazolio (MTT; Sigma-Aldrich) method according to the manufacturer’s instructions. Optic densities were measured at 560/620 nm in a Multiskan Reader MCC/340 Labsystems. Three independent experiments with triplicate measurements were performed. Evaluation of inhibition of CPE using the MTT method was done for two of the three experiments. Data were analyzed by a four-parameter curve-fitting from a dose-response curve using GraphPad Prism (version 7.00) to calculate the EC_50_ (concentration of the compound that inhibited 50% of the infection) based on visual CPE scoring or based on the MTT method.

### Assessment of Cell Viability

To assess the effects of the compounds on Caco–2 cell viability, cell viability was measured in confluent cell layers treated with a range of compound concentrations in absence of virus using the Rotitest Vital (Roth) according to manufacturer’s instructions, as previously described (*12*). All assays were performed three times independently in triplicate. From this the CC_50_ (cytotoxic concentration of the compound that reduced cell viability to 50%) was calculated from a dose-response curve in GraphPad Prism (version 7.00) using four-parameter curve-fitting.

### Selectivity Index

The selectivity index for each of the compounds was determined as the ratio of the CC_50_ to the EC_50_.

### Modelling

In general, *in silico* docking can be a useful approach for identifying subsets of molecules for *in vitro* studies and can be used to explain *in vitro* observations on a structural level. The coordinates of the crystal structure of the main SARS-CoV-2 protease were retrieved from the PDB database (https://www.rcsb.org/structure/6lu7).

Preparing DRV and the protein for *in silico* molecular docking was performed with the software package MOE (Molecular Operating Environment 2019.01; Chemical Computing Group ULC, Montreal, QC, Canada, H3A 2R7, 2019). The force field used was AMBER ETH:10 with default ‘quickprep’ settings to prepare the protein complex. The original ligand was then removed. General docking settings were then altered to have 50 initial placements.

## Results

### Assessment of *In Vitro* Antiviral Activity, Cytotoxicity and Selectivity

Remdesivir showed strong antiviral activity against SARS-CoV-2 with an EC50 of 0.11 µM based on visual scoring of inhibition of CPE (Figure 1a). In the same experiments, DRV did not show any inhibition of SARS-CoV-2 induced CPE (Figure 1b; EC_50_ >100 μM). Similar results were obtained using the MTT method. Remdesivir showed potent antiviral activity with an EC_50_ value of 0.38 μM (Figure 1a), while DRV showed no effect (EC_50_ >100 μM, Figure 1b). No cytotoxicity of remdesivir or DRV was observed on Caco-2 cells with CC_50_ values >100 μM. The selectivity index (CC_50_ / EC_50_) for DRV could not be calculated (>100 µM / >100 µM for SARS-CoV-2) due to the lack of antiviral activity. In contrast, remdesivir had a selectivity index of >900 by visual CPE scoring and >260 by the MTT method, confirming a strong *in vitro* antiviral effect against SARS-CoV-2.

**Figure 1.**
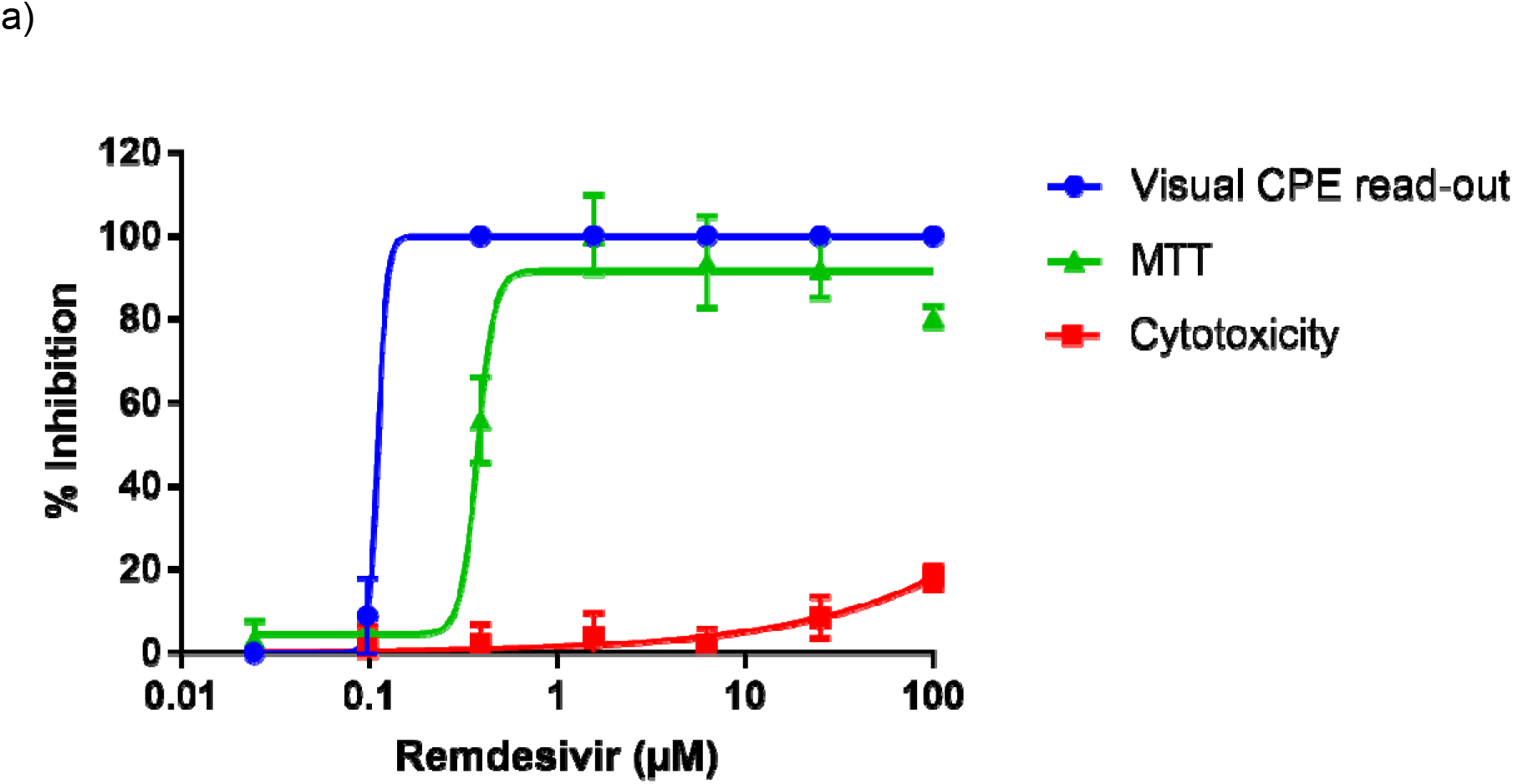
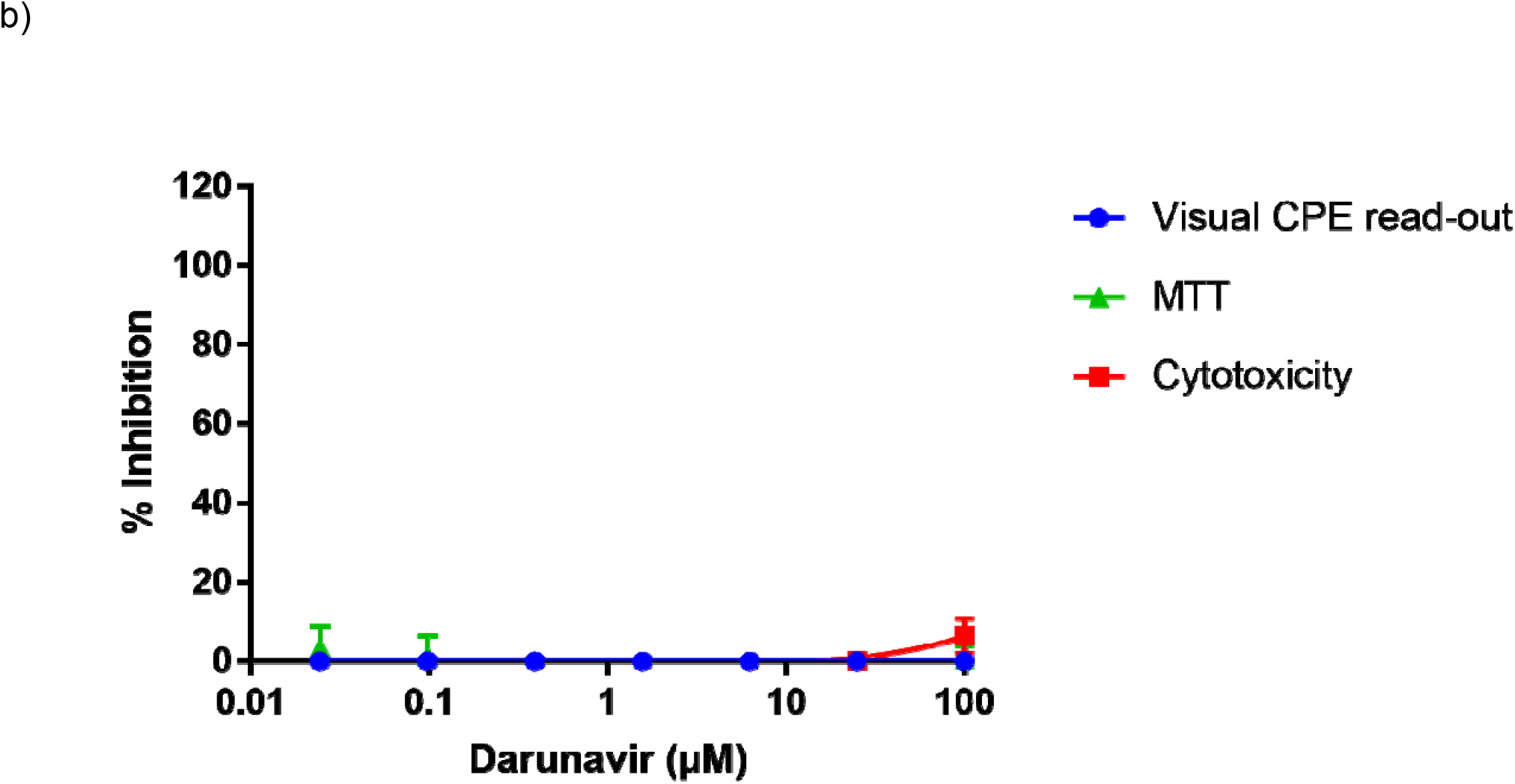
Inhibition of SARS-CoV-2 in Caco-2 cells by cytopathogenic effect assay using a visual read-out (visual CPE read-out) and MTT assay (MTT) following addition of a) remdesivir, or b) darunavir (DRV). Cytotoxicity data (measured by MTT on uninfected cells) are also shown. Mean percent inhibition for each readout across three independent experiments with triplicate measurements are plotted (2 independent experiments for the MTT assay). The error bars represent the standard deviation. Visual CPE-read out EC_50_ = 0.11 μM; MTT EC_50_ = 0.38 μM; CC_50_ >100 μM; Selectivity index = >900 (Visual CPE read-out); >260 (MTT) Visual CPE read-out EC_50_ > 100 μM; MTT EC_50_ > 100 μM; CC_50_ >100 μM

### Modelling

*In silico* docking of DRV in the crystal structure of the SARS-CoV-2 main protease (3CL protease) identified five docking poses. The docking scores ranged from S= −8.6 to −8.2, showing that DRV can fit into the pocket, but these values are indicative of suboptimal binding to this protein. Visual inspection of each of these poses showed very few interactions of DRV with the active site of the protease, and the catalytic cysteine residue was not directly targeted, unlike the many strong interactions observed for DRV bound to the HIV protease (14).

## Discussion

Current efforts to manage the COVID-19 pandemic have largely focused on improved hygiene, quarantine of infected individuals, social distancing to limit transmission and development of a vaccine (*15*). Despite the expedited efforts to develop a vaccine and collaborative efforts to screen compounds in discovery and development across the broader pharmaceutical industry for activity against COVID-19, patients are in immediate need of therapeutic interventions (*12, 16*).

Current data on the therapeutic effect of HIV protease inhibitors in patients with COVID-19 are far from comprehensive. This study demonstrated that DRV showed no *in vitro* antiviral activity against SARS-CoV-2 at clinically relevant concentrations. Furthermore, structural analyses using protease structures are consistent with these data. DRV binds to the active site of the HIV virus’ dimeric aspartic protease (*14*). The crystal structure of this protease is well-elucidated and has been shown to have an extensive hydrogen-bonding network with DRV, allowing for the potent *in vitro* activity (EC_50_ values = 1.2 to 8.5 nM) of this protease inhibitor against HIV (*6-7*). By contrast, the SARS-CoV-2 main protease is a cysteine protease (Protein Data Bank-code 6LU7) and while several docking poses have been found for DRV in *in silico* models, unlike in HIV, these poses showed little interaction with the SARS-CoV-2 main protease active site. Several publications describe *in silico* docking experiments on the main coronavirus protease that specifically focus on or include DRV (*17–21*). Although these studies suggest DRV as a candidate for further investigation, such promising docking results could not be reproduced in our *in silico* docking studies. Such discrepancies can often result from *in silico* docking, which is primarily a useful approach for identifying subsets of molecules for *in vitro* activity testing. No *in vitro* antiviral activity of DRV against SARS-CoV-2 was found in the experiments reported here. In this study, remdesivir demonstrated activity against SARS-CoV-2 with an EC_50_ of 0.38 μM, which is in line with the earlier reported remdesivir EC_50_ of 0.77 µM, indicating that the *in vitro* antiviral assay used is appropriate to assess antiviral activity against SARS-CoV-2 (*12*).

In conclusion, the lack of *in vitro* antiviral activity of DRV against SARS-CoV-2 does not support the use of DRV for treatment of COVID-19. Hence, the use of DRV (boosted with either ritonavir or cobicistat) should remain solely for treatment of patients with HIV infection.

## Data Availability

The data sharing policy of Janssen Pharmaceutical Companies of Johnson & Johnson is available at https://www.janssen.com/clinical-trials/transparency. As noted on this site, requests for access to the study data can be submitted through Yale Open Data Access (YODA) Project site at http://yoda.yale.edu.

## Acknowledgments

Medical writing supporting for the development of this manuscript was provided by Patrick Hoggard of Zoetic Science, an Ashfield company, part of UDG Healthcare plc; this support was funded by Janssen Pharmaceuticals. We thank Lena Stegmann for technical support by antiviral assays.

## Disclaimer

Sandra De Meyer, Christophe Buyck, Ellen Van Damme, Marnix Van Loock and Brian Woodfall are employees and may be stock owners of Johnson and Johnson. Sandra Ciesek received research funding from Janssen for this research.

Funding for this study was provided by Janssen Pharmaceutica.

